# Economic evaluation of disease elimination: an extension to the net-benefit framework and application to human African trypanosomiasis

**DOI:** 10.1101/2021.02.10.20181974

**Authors:** Marina Antillon, Ching-I Huang, Kat S Rock, Fabrizio Tediosi

**Affiliations:** Swiss Tropical and Public Health Institute, Basel, Switzerland; University of Basel, Basel, Switzerland; Zeeman Institute, University of Warwick, Coventry, UK; Mathematics Institute, University of Warwick, Coventry, UK

**Keywords:** eradication, elimination, economic evaluation, mathematical modeling

## Abstract

The global health community has earmarked a number of diseases for elimination or eradication, and these goals have often been praised on the premise of long-run cost-savings. However, decision-makers must contend with a multitude of demands on health budgets in the short- or medium-term, and costs-per-case often rise as the burden of a disease falls, rendering such efforts beyond the cost-effective use of scarce resources. In addition, these decisions must be made in the presence of substantial uncertainty regarding the feasibility and costs of elimination or eradication efforts. Therefore, analytical frameworks are necessary to consider the additional effort for reaching global goals, like elimination or eradication, that are beyond the cost-effective use of country resources. We propose a modification to the net-benefit framework to consider the implications of switching from an optimal strategy, in terms of cost-per-burden-averted, to a strategy with a higher likelihood of meeting the global target of elimination of transmission by a specified date. We illustrate the properties of our framework by considering the economic case of efforts to eliminate transmission of *gambiense* human African trypanosomiasis (gHAT), a vector-borne parasitic disease in West and Central Africa, by 2030.

**Significance Statement:** Various diseases are earmarked for elimination by the global health community. While the health economic implications of elimination have been discussed before, the combination of uncertainty, cost-effectiveness in terms of cases averted, and elimination in the face of rising per-case costs has not been tackled before. We propose an approach that considers the tension between the dual objectives of cost-effectiveness and elimination while incorporating uncertainty in these objectives. We apply our method to strategies against human African trypanosomiasis in three settings, but this method could be directly applied to simulation-based studies of the cost-effectiveness of other disease elimination efforts. The method yields common metrics of efficiency when stakeholders have different objectives.

---

**T**he successful eradication campaigns of smallpox and rinderpest have curried political support for the elimination or eradication of transmission (EEOT) of other diseases. Thus far, regional elimination has been achieved for land-transmitted rabies in Europe and for malaria in large parts of the world. Yet, due to the strong correlation between many of the diseases targeted for EEOT and poor sanitation, health infrastructure, or overall material conditions, such disease campaigns are disproportionately focused in low-resource settings, bringing to the fore important questions about the efficiency of EEOT efforts (1, 2).

On the one hand, expensive EEOT interventions are often justified on the basis of future cessation of activities; one front-loads the expenses on a disease on the premise that public health activities can cease in the not-too-distant future, at which time investments will be recovered. Smallpox eradication is claimed to have saved, within just a few years, billions of dollars (3). However, falling just short of EEOT could be the worst of all possible scenarios: one has diverted ever-increasing resources from other purposes, but one cannot cease activities and recover investments. In part because of the risk of failure and in part because of the accelerating per-case cost of campaigns near the end-game, such efforts might not be considered an efficient use of resources from the perspective of decision-makers with limited budgets contending with a variety of health challenges. For instance, there were 22–176 wild-type poliomyelitis cases reported yearly between 2016–2020 but eradication campaigns cost approximately $1B due to the continued need for surveillance and vaccination globally (4, 5). Guinea worm disease (GWD) caused only 54 cases in 2019, yet it is the subject of campaigns that cost approximately $30M (6, 7). The eradication targets of both diseases have experienced delays of 23 years and 10 years, respectively, therefore stalling the promise to recover investments (7, 8).

To explore, scrutinize, and provide insights into questions surrounding EEOT, the epidemiology and health economics fields have a rich toolbox that captures the non-linear transmission dynamics, temporal features, and economic implications of disease control. Separately, a few studies have tried to grapple with questions around the economic implications of EEOT by employing game theoretic approaches (3, 9, 10). These studies have designated different levels of coverage (i.e. vaccine coverage) necessary for “control” and EEOT, and the difference in costs between “control” and EEOT strategies constitutes the price for elimination (3, 9, 10). While making the problem theoretically tractable, the application is quite narrow: in practice, addressing the burden of a disease may require various distinct activities, and parsing the activities that contribute uniquely to “control” or EEOT is not always possible, as each activity contributes to both objectives to varying degrees.

Moreover, there is a lot of uncertainty in whether strategies lead to EEOT. In practice one cannot purchase EEOT with certainty, as GWD and polio have shown, one merely invests in activities that are conducive to EEOT, and therefore, the absence of probabilistic thinking in previous literature fails to capture a key component of the decision-making process. Frameworks with multiple objectives are exceedingly rare, and while some work has been done to consider policy analyses of stakeholders with differing fundamental objectives within a modelling framework (i.e. Probert *et al*. (11)), such approaches have seldom been developed for cost-effectiveness analyses (12) and never for analyses taking into account EEOT objectives.

Here we develop a framework that can handle 1) strategies that have different probabilities of EEOT, 2) where activities are not easily classified as exclusively “control” or “elimination” activities, 3) and where multiple objectives – specifically cost-per-DALY-averted and EEOT – are transparently considered. We extend the net-benefit framework, useful for decision-analysis in the presence of uncertainty, in order to evaluate cost-effectiveness of public health strategies while explicitly outlining the ‘premium’ of elimination, or the additional resources that are necessary to bring a country’s activities in line with global goals. One important feature of our framework is that it is operationalized within a probabilistic simulation framework, which makes it a simple extension to ubiquitously used approaches for decision analysis in the face of uncertainty, thereby being applicable in a wide array of situations.

We then apply our new framework to *gambiense* human African trypanosomiasis (gHAT) in three distinct regions of the Democratic Republic of Congo (DRC). The three regions highlight the strengths of our framework and its applications under different circumstances: circumstances of certainty, uncertainty, and where more than two strategies are reasonable candidates to abate disease burden and interrupt disease transmission.

## Background

Our objective is to introduce an extension of the net-benefit framework to account for the resource implications of aligning potentially incongruous objectives of efficiency and EOT.

The keystone metric of value-for-money in cost-effectiveness analysis is the incremental cost-effectiveness ratio (ICER), which we touch on briefly as a point of departure for the net-benefit framework. The ICER is defined as the ratio of the difference in costs, Δ*C*, and the difference in health effects, Δ*E* of two interventions:

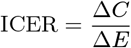

where the change in costs and health effects are computed as the net difference in costs and effects between strategies. Effects are usually denominated in disability-adjusted life-years (DALYs), a metric that is comparable across diseases. For the purpose of our analysis, the effects will be distinguished between DALYs and the probability of elimination of transmission (EOT).

Between two strategies, a second strategy will be considered cost-effective compared to the first (the comparator strategy) if the ICER does not exceed a health planner’s willingness-to-pay (*λ*^WTP^) per DALY averted (13, 14).

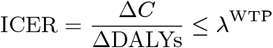

The health planner’s *λ*^WTP^ per DALY averted is equivalent to the ICER of the least efficient strategy (the strategy with the highest ICER) in the health portfolio (15–18). Recently the World Health Organization (WHO) has advocated that cost-effectiveness results should be shown at a range of WTP values; Table 1 describes select values in the context of DRC, the country that will be the subject of our examples.

**Table 1.** **Contextualizing willingness-to-pay values 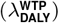 for low-income settings**.

| $\lambda_{\text{DALY}}^{\text{WTP}}$ | Rationale |
| --- | --- |
| \$0 | This is cost-saving or cost-neutral over the chosen time horizon of the analysis. It should be noted that annual expenditure or budgets not necessarily static across the whole period for all (or any) strategies. |
| \$250 | Two studies that modelled the real investments made across countries estimated that the investments in DRC are \$5–\$230 per DALY averted in 2013 US\$ (18) or \$54–\$69 per DALY averted in 2015 US\$ (19). We rounded up to \$250 for convenience. |
| \$500 | Approximately equivalent to the annual gross national income (GDP) of DRC in 2018, which was the definition of a “very cost-effective” strategy as delineated in the WHO CHOICE program (20). |
| \$1500 | Approximately equivalent to three times the annual gross domestic product (GDP) of DRC, which was the definition of a “cost-effective” strategy as delineated in the WHO CHOICE program (20). |

Not surprisingly, uncertainty in costs and DALYs exists, in particular with regards to diseases in populations that are difficult to study or diseases that have been historically neglected. There exists a literature devoted to the difficulties of accounting for parameter uncertainty when calculating ICERs while remaining consistent with the economic principles on which cost-effectiveness is grounded, primarily because ICERs are ratios (13, 21, 22). The net-benefit framework was therefore developed to circumvent some of the issues surrounding ICERs and uncertainty. The NMB is a simple arithmetic rearrangement of the ICER into a linear additive formulation:

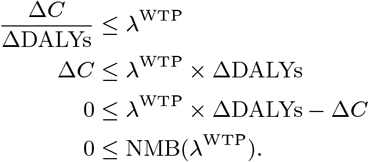

where *λ*^WTP^ is the a monetary value for DALYs avoided. The linear additive formulation circumvents the technical issues with samples of ratios (22). Given a Monte Carlo sample of *N* iterates of the disease and cost model, a strategy is preferred over the comparator if the expected NMB exceeds zero:

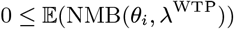

where *θ* is a the parameter vector and *I* ∈ {1, …, *N*} are the iterates of each parameter. In a multiple-strategy decision analysis between *J* interventions, the preferred strategy is the strategy that maximizes 𝔼 (NMB(*θ*_*i*_, *λ*^WTP^)):

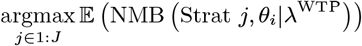

the differences refer to the difference between the comparator strategy (*j* = 1) and any alternative strategy *j* ∈ 2 : *J* in the analysis.

Simultaneously, the framework allows for a probabilistic interpretation of cost-effectiveness by conditioning on *λ*^WTP^:

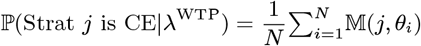

Where

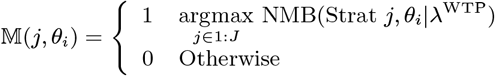

The algorithm therefore presents a measure of certainty that the strategy with the highest expected NMB is optimal over all other strategies, given by the proportion of samples where the strategy has the highest NMB of all strategies (13, 23).

The value judgements implied here are quite simple: the only relevant values are net costs (including the benefit of cost savings for the program by averted disease) and DALYs averted. Other considerations not accounted for include: the socioeconomic distribution of the beneficiaries vs remaining disease victims, ethical considerations, or if a pivotal threshold for disease evolution, such as re-emergence or eradication can be reached (24).

### Economic evaluation framework for multiple objectives

Our proposal makes explicit the relationship between the WTP of dual objectives for averting disease burden, denominated in DALYs averted, and elimination, while taking into account the uncertainty in achieving those objectives and the concomitant costs.

Net Monetary and Elimination Benefits (NMEB(*θ*_*i*_))

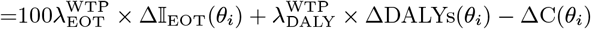

where the indicator function, Δ𝕀_EOT_ is 1 if EOT is achieved by only one strategy and 0 if neither or both strategies achieve EOT (for parameter set *θ*_*i*_ and strategy *j*):

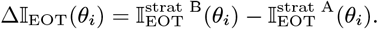

The use of 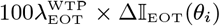 incorporates the WTP to raise the probability of elimination. While the term 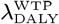 is interpreted as the highest price paid to avert an additional DALY, 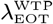 is interpreted as highest price paid per additional point increase in the probability of elimination.

The linear additive scale allows us to decompose the resource expenditure into two portions: the portion that is justifiable based on disease burden averted and the portion of the expenditure that is justifiable by concerns about global goals. While elimination and disease control are not separable or independent, which is accounted for in the dynamic transmission model, the linear form in this formulations allows us to separate the elimination benefits related to disease control (decreasing DALYs) and all other benefits of elimination above and beyond averted disease burden. This circumvents the need to calculate *C*_DALY_ and *C*_EOT_, which are rarely separable as no activity will contribute to a reduction in one metric without impacting the other.

In a manner analogous to the traditional NMB, the strategy that ought to be implemented is indicated by:

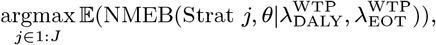

and the probability that a strategy is cost-effective is

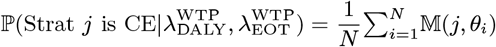

Where

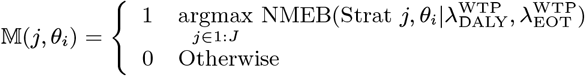

### Premium of elimination

A useful metric easily calculated from the formulation of NMEB is the *premium of elimination*. We begin with a simple context: if we assume there are two strategies, which are to reach elimination with 0% and 100% certainty respectively, and we suppose that the elimination strategy would not avert any additional DALYs over the non-elimination strategy (e.g. detection and treatment are superb), then the expected premium of elimination, given here by Premium_EOT_, would equal the expected cost difference between the two strategies:

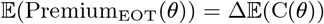

However, in practice any two or more strategies are unlikely to avert the same number of DALYs; in fact, most often the elimination-prone strategy is likely to avert some additional DALYs, albeit at a potentially high cost. The health planner has a certain 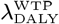 for those additional DALYs that the elimination strategy averts, though perhaps not a WTP that would completely bridge the gap in costs between the two strategies. In such a case, the expected Premium_EOT_, is:

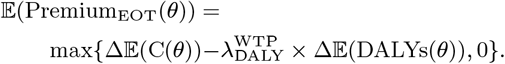

In other words, the more the health planner is willing to pay for DALYs averted, the lower the additional premium that will be paid for elimination.

If one strategy has both a higher probability of achieving the elimination and a relatively low incremental cost, then the premium of elimination is equal to zero, as no additional resources are needed to justify elimination beyond those resources traditionally considered to be efficient (cost-effective) to avert disease.

### Premium of elimination and NMEB

The NMEB and the Premium_EOT_ are linked as follows.

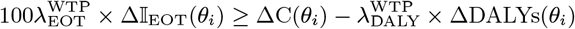

taking the expectation of both sides of the equations:

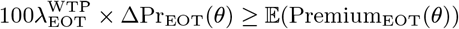

In other words, the premium of elimination must be smaller than the product of the between-strategy differential probability of EOT and the WTP for additional certainty of EOT. For instance, if the comparator strategy (strategy A) is the preferred strategy under the traditional NMB framework, then in order to select strategy B on the basis of its higher probability of elimination, the product of 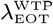 and the differential probability must be at least as large as the Premium_EOT_.

### Premium of elimination, discount rates, and time horizons

Inadequately chosen time horizons and discount rates would impact the size of the premium of elimination, presenting not the monetary value of additional benefits of elimination, but rather uncaptured savings. While the benefits of elimination or eradication could reach infinitely into the future, one does not generally include these infinite benefits or savings for two reasons. First, the principles of time-preference generally dictate that we apply a discount rate, which would make the costs and benefits more than two or three decades into the future worth almost zero in present-day terms. Even if no discounting were to be taken into account, most analyses of interventions of non-chronic infectious diseases have time horizons under 20 years, and picking a longer time horizon for our analysis would not allow comparability across diseases in a health planner’s portfolio (20). Lastly, it is recognized that lifetime rewards for current-day strategies against infectious diseases are difficult to incorporate as the state of epidemics several decades into the future can be influenced by factors that cannot be adequately predicted (25). Therefore, we caution against overly short time horizons or unusually high discount rates, as this could raise the premium of elimination (by the same token, unusually long time-horizons and low discount rates would lower the premium of elimination).

## Results

### Health outcomes, costs, and traditional ICERs

Using a joint transmission and cost model we made projections of the epidemiological impact the resource use over 2020–2040 of four strategies in three locations against gHAT (Table 2 provides an overview on the component interventions, while further details can be found in the Materials and Methods). In Region 1, success or failure of the 2030 elimination of transmission (EOT) goal is certain depending on the selected strategy, but in Regions 2 and 3 success and failure of the EOT goal is uncertain (Table 3).

**Table 2.**
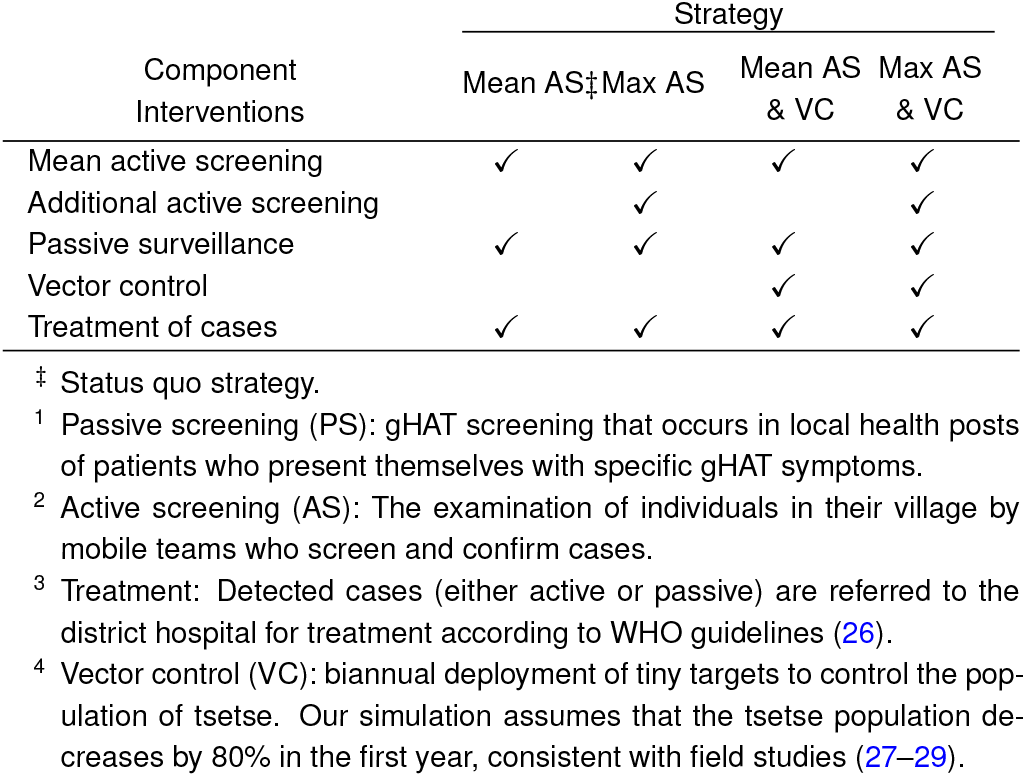
**Strategies for control and elimination of gHAT in a typical endemic health district**.

| Component Interventions | Strategy |  |  |  |
| --- | --- | --- | --- | --- |
|  | Mean AS<sup>‡</sup> | Max AS | Mean AS & VC | Max AS & VC |
| Mean active screening | ✓ | ✓ | ✓ | ✓ |
| Additional active screening |  | ✓ |  | ✓ |
| Passive surveillance | ✓ | ✓ | ✓ | ✓ |
| Vector control |  |  | ✓ | ✓ |
| Treatment of cases | ✓ | ✓ | ✓ | ✓ |

<sup>‡</sup> Status quo strategy.
<sup>1</sup> Passive screening (PS): gHAT screening that occurs in local health posts of patients who present themselves with specific gHAT symptoms.
<sup>2</sup> Active screening (AS): The examination of individuals in their village by mobile teams who screen and confirm cases.
<sup>3</sup> Treatment: Detected cases (either active or passive) are referred to the district hospital for treatment according to WHO guidelines (26).
<sup>4</sup> Vector control (VC): biannual deployment of tiny targets to control the population of tsetse. Our simulation assumes that the tsetse population decreases by 80% in the first year, consistent with field studies (27–29).

**Table 3.**
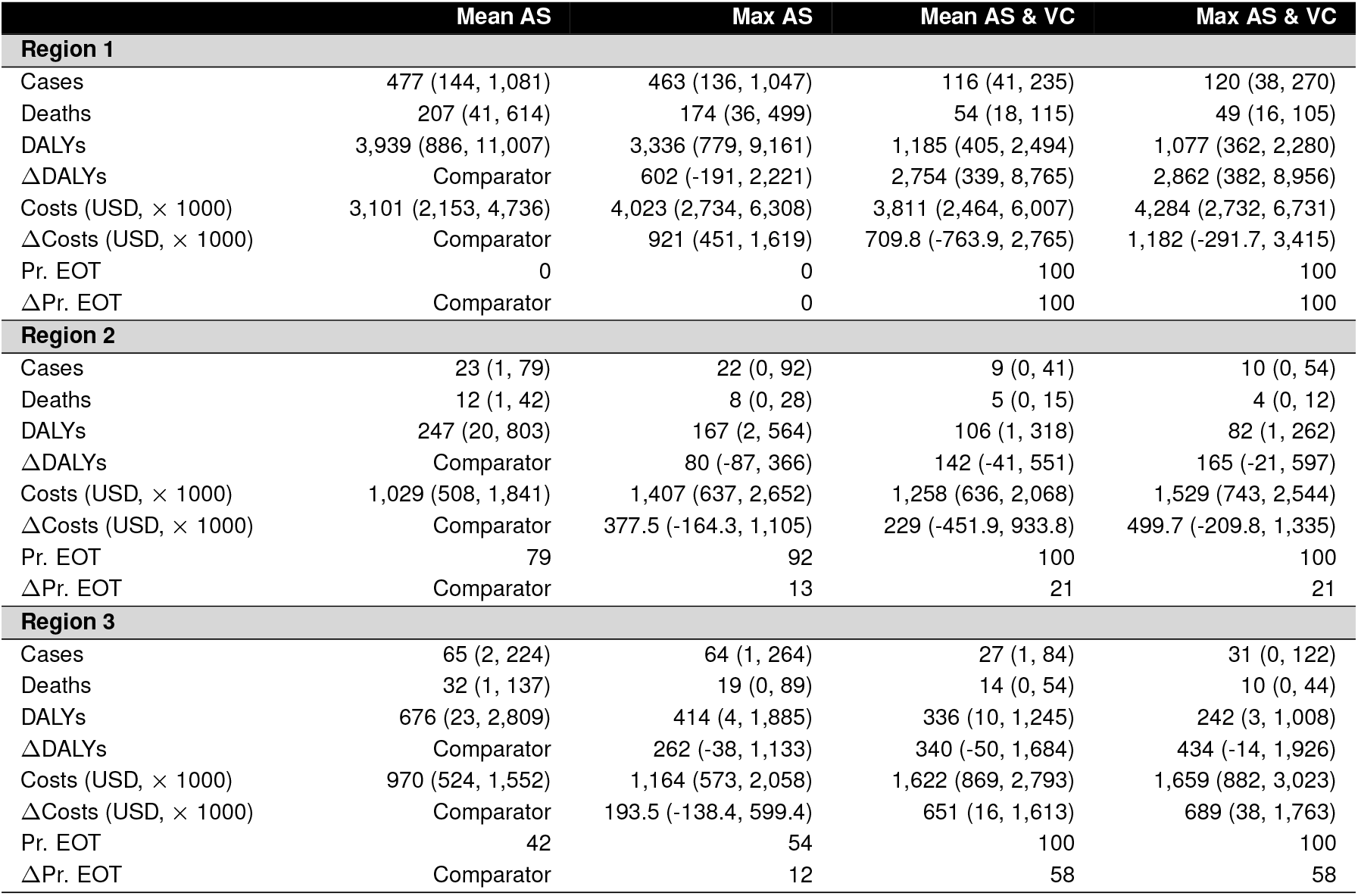
**Intermediate outcomes, cost-effectiveness, and elimination of transmission in three sample health zones**.

If the status (comparator) strategy remains in place (Mean active screening, AS) there will be an average of 477 cases and 207 deaths in Region 1, 23 cases and 12 deaths in Region 2, and 65 cases and 32 deaths in Region 3. In terms of DALYs, those incidence levels give rise to 3,939 DALYs in Region 1, 247 DALYs in Region 2, and 676 DALYs in Region 3. Under any strategy in all settings, the burden of disease is expected to decline, but strategies with VC are expected to expedite this decline substantially (see Fig S3).

Costs per year show that while strategies that include vector control (VC) are more costly in the short-run, the investments begin to yield returns after 2028 in Region 1, after 2025 in Region 2, and after 2030 in Region 3 (Fig S4). Costs are driven by AS activities, and when applicable, by VC activities, so the timing of cessation of these activities plays an important role in the ability of ambitious investments to be recovered (see Fig S5). Nevertheless, it is not certain that these strategies will yield cost-savings over a 20-year investment period, although total costs are only marginally higher compared to the comparator (Mean AS).

### Net Monetary Elimination Benefits: where success and failure are certain

The probability of EOT in Region 1 is shown in 1A and the results of our decision analysis under the traditional net benefits framework is shown in Figure 1B. After taking into account parameter uncertainty, our analysis shows that Mean AS has an 80% probability of having the minimum cost (optimal at 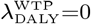). However, if 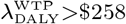, the strategy Mean AS & VC is optimal with 47–55% probability.

**Fig. 1.**
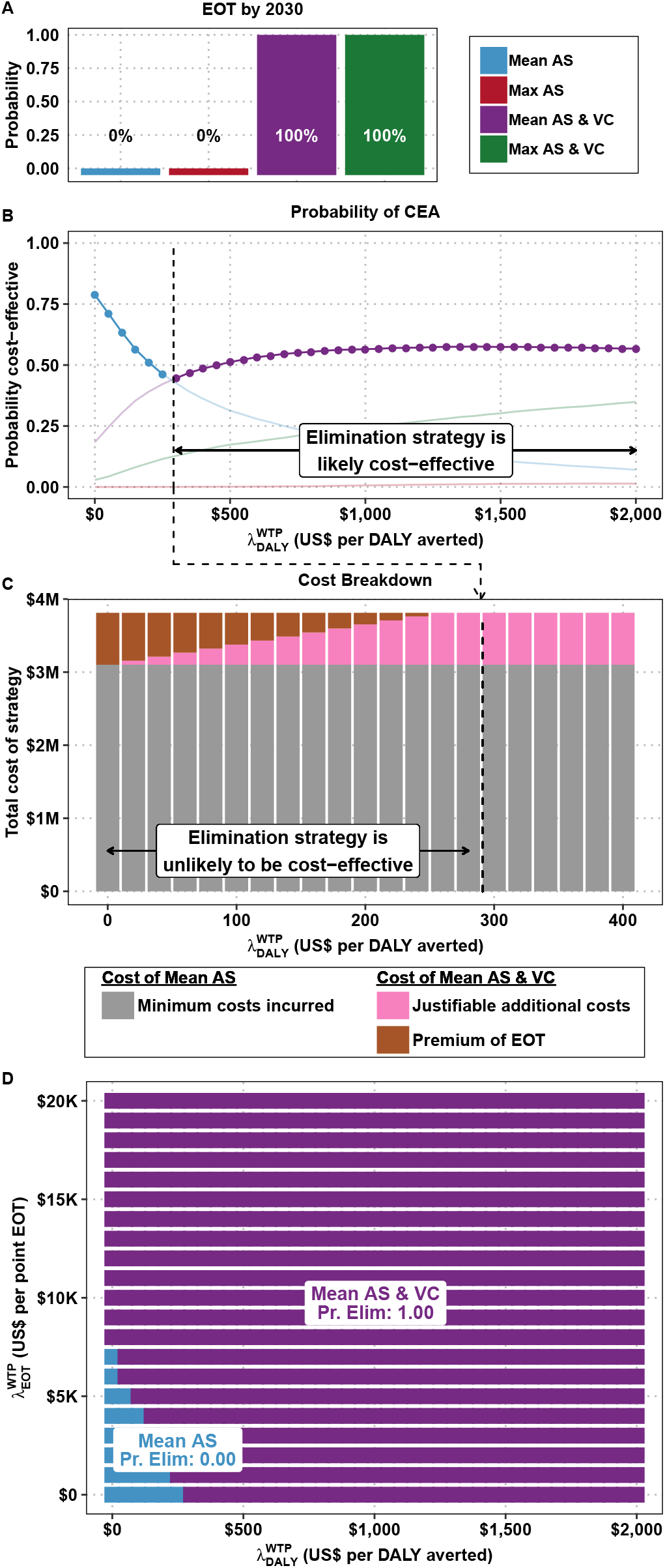
Cost-effectiveness acceptability curves and cost breakdown for Region 1. A) the probability of EOT by 2030 for each strategy. B) The traditional cost-effectiveness acceptability frontiers (CEAFs). C) The total cost for a strategy that reaches elimination with 100% probability and the breakdown between minimum cost strategy, and justifiably additional costs and premium of elimination across 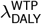 values. D) Cost-effectiveness acceptability heatmaps (CEAH).

In Region 1, Mean AS has a 0% probability of EOT by 2030, whereas Mean AS & VC has 100% probability of EOT. The expected Premium_EOT_, is shown in 1C. In a policy environment of low 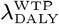, any health planner must be able to justify the entire $709,800 cost difference between Mean AS and Mean AS & VC on the basis of EOT alone. When DALYs are not valued monetarily 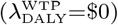, the Premium_EOT_ is simply equivalent to the difference in costs between the two strategies. If, for instance, 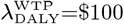, a health planner must be able to justify only a Premium_EOT_ of $434,400, as the other $275,400 that would be justifiable for DALYs averted. In a policy environment of a 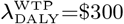, the strategy that reaches elimination is entirely justifiable on the health gains achieved (DALYs averted), and the Premium_EOT_ is therefore $0.

Fig 1C shows the optimal choice of strategy for a range of 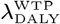 and 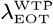 values. In a policy environment where 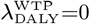 and 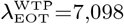 per additional probability point of EOT, the optimal strategy guarantees elimination, as that is the 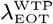 that justifies the $709,800 premium of elimination.

### Net Monetary Elimination Benefits: where success and failure are uncertain

Region 2 illustrates a setting in where the comparator strategy (Mean AS) has a 79% probability of EOT, and therefore the binary conception of “control” or “elimination” strategies fails to adequately capture the decision-maker’s dilemma.

The conditions for which an economically optimal strategy maximizes the probability of EOT is shown in Fig 2. If 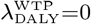, a strategy that almost ensures EOT – Mean AS & VC – is optimal if 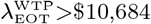 per additional percentage point, and the Premium_EOT_ is $229,000 (Table S3). If, however, 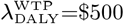, then the Premium_EOT_ is $158,000 and the 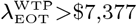. At 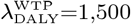, then Premium_EOT_ decreases to $16,000 and 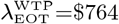.

**Fig. 2.**
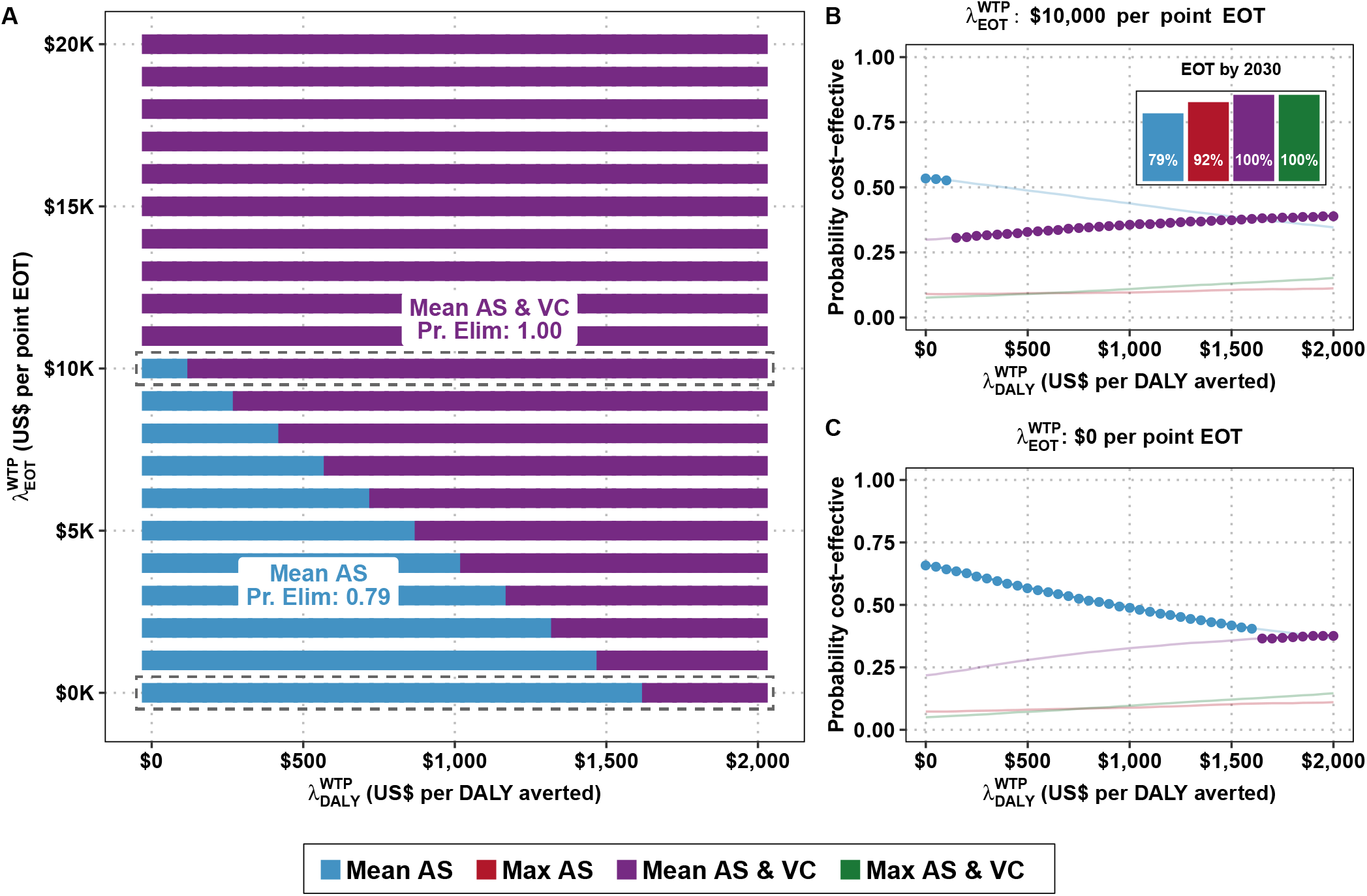
Cost-effectiveness acceptability heatmaps for Region 2. A) Cost-effectiveness acceptability heatmap (CEAH). Along the x-axis is the cost-effectiveness threshold for averting disease burden, 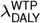, and along y-axis is the cost-effectiveness threshold for elimination of transmission (EOT), 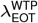, to raise the probability of EOT by 2030 by one percentage point. B–C) Cost-effectiveness acceptability frontiers (CEAFs). The inset in the top-right graph is the probability of each strategy’s EOT by 2030.

Our most complex setting, Region 3, is shown in Fig 3. Under the traditional net benefits framework, either the Mean AS or Max AS strategy are cost-effective at 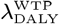 values consistent with historical investment levels in low-income countries (Fig 3C), but these strategies have only a 42% and 54% probability of EOT (Fig 3B). Without an investment in elimination justified by benefits that extend beyond the averted DALYs, achievement of EOT is uncertain.

**Fig. 3.**
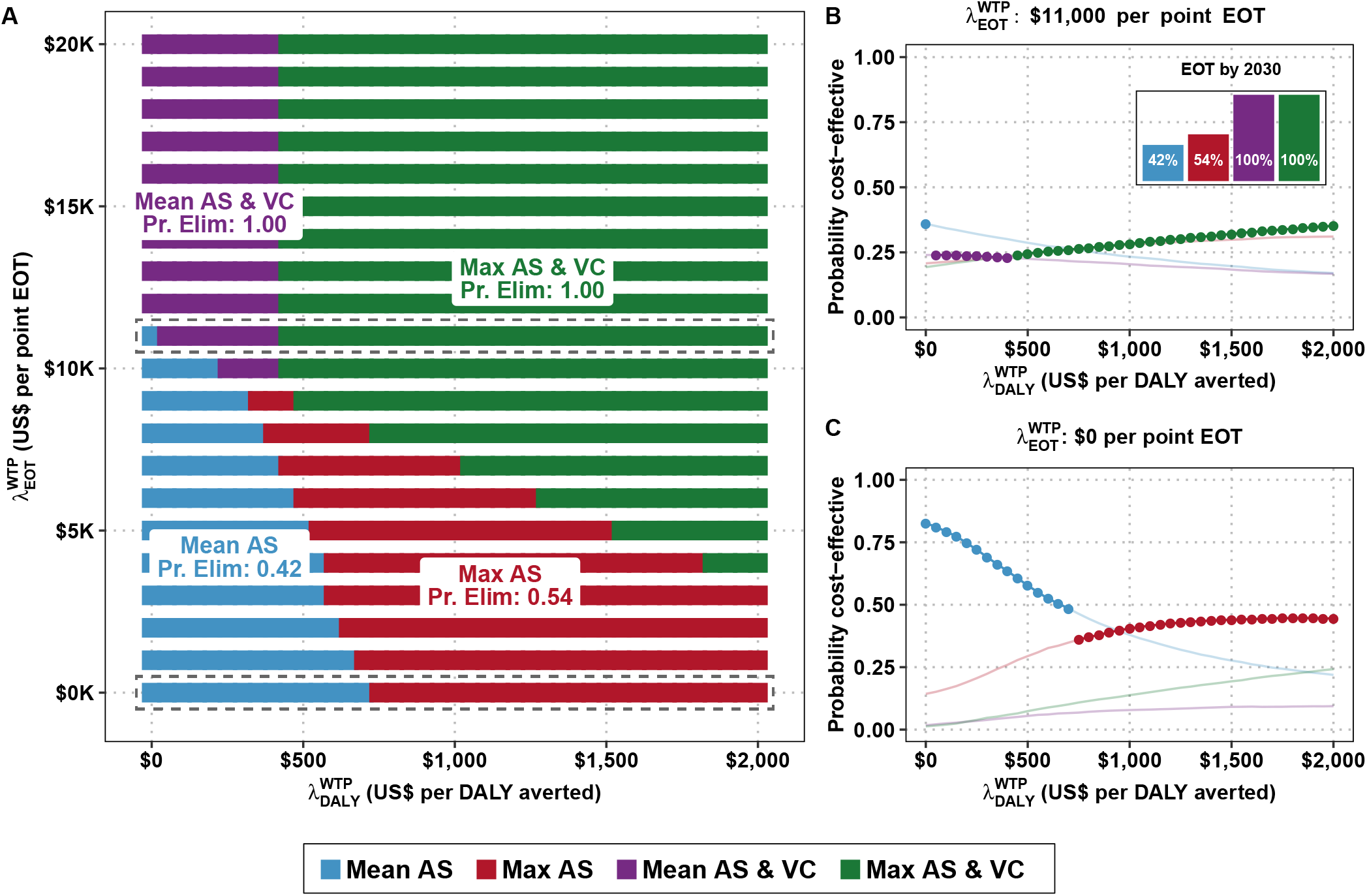
Cost-effectiveness acceptability heatmaps for Region 3. A) Cost-effectiveness acceptability heatmap (CEAH). Along the x-axis is the cost-effectiveness threshold for averting disease burden, 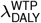, and along y-axis is the cost-effectiveness threshold for elimination of transmission (EOT), 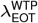, to raise the probability of EOT by 2030 by one percentage point. B–C) Traditional cost-effectiveness acceptability frontiers (CEAFs). The inset in the top-right graph is the probability of each strategy’s EOT by 2030.

At 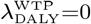, then a decision-maker must have a 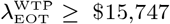 and incur a Premium_EOT_=$194,000 to bolster the chances of elimination from 42% to 54% or 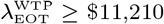 and incur a Premium_EOT_=$651,000 to bolster the probability of elimination from 42% to >99%. Therefore, as long as one does not value DALYs averted, the switch from Mean AS to Max AS would incur a lower premium, but switching to Mean AS & VC would be more efficient on the basis of per-point probability of reaching EOT (Fig 4 and Table S4).

**Fig. 4.**
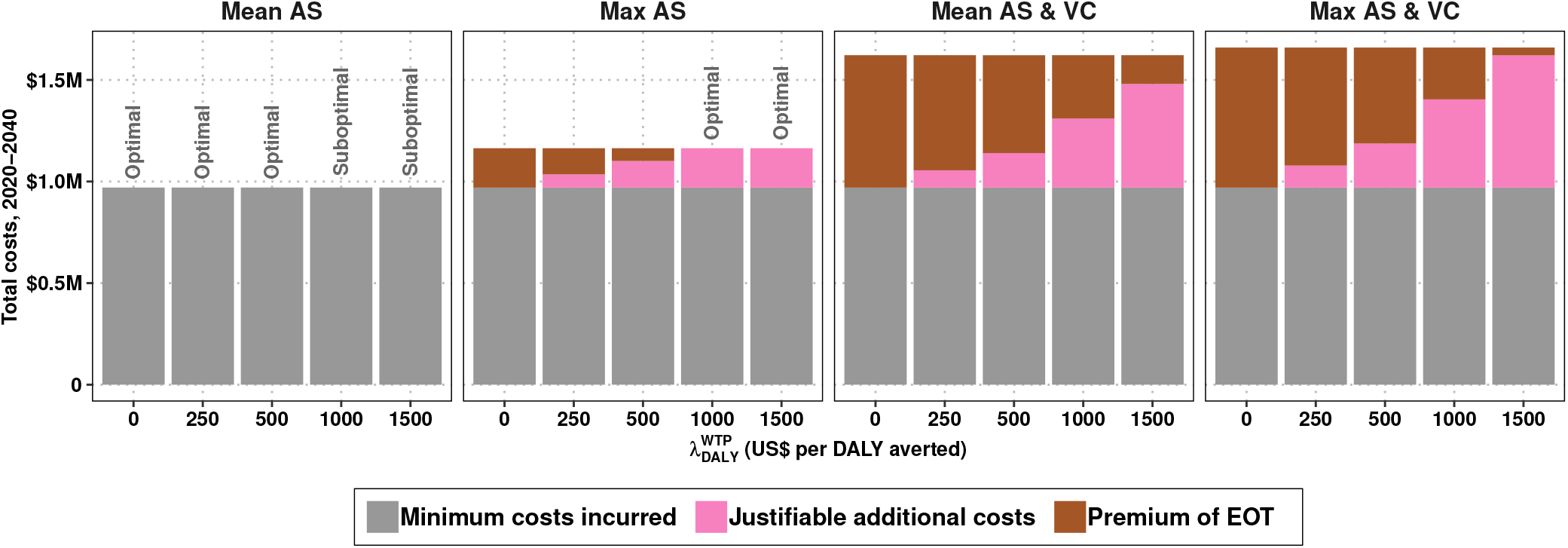
Premium of elimination in Region 3, across different values of 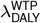, contextualized in table 1.

If, however, the policy environment is one where 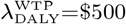, then Mean AS & VC has become dominated by both objectives: Max AS will avert more DALYs than Mean AS & VC because it treats more extant infections in the short-term, and Max AS & VC is more efficient at both averting DALYs and raising the probability of EOT on a per-point basis. To maximise the probability of EOT when 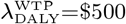, one would deploy Max AS & VC, which has a Premium_EOT_=$472,000, equivalent to 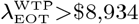. At 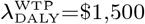, the strategy Max AS is cost-effective, so the Premium_EOT_ of Max AS & VC is $237,000 and 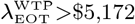.

## Discussion

Much of the early literature about cost-effectiveness and decision-making in the presence of uncertainty was about analytical methods to do what is now more easily done with simulation (14, 21, 30, 31). Although simulation of a wide array of epidemiological and policy questions is ubiquitous, the interpretation of those simulations must rest within frameworks which decompose uncertainty and frame economic optimization in a manner that is in line with economic theory. The existing tools in the health economics toolbox generally answer the question “are public health efforts economically justified?” but when the global health community has set ambitious goals, such as elimination of infectious diseases, questions of resource use across different administrative levels (local, national, regional, and global) do not fit neatly in existing frameworks and one asks “to what degree are public health objectives economically justified by various objectives?” In the context of EEOT of infectious diseases, there may be a tension between EEOT objectives and maintaining an efficient use of scarce resources (32). This is particularly salient because the same resources – in-kind, capital, and financial – that vie for EEOT efforts could be diverted to address other urgent health goals.

We have presented an extension to the net benefits framework to inform decisions that contain an elimination objective that may stand at odds with concerns about efficient resource allocation. Our proposed framework explicitly models the additional premium for elimination activities in the presence of uncertainty.

The illustrative analysis shows that in Region 1, EOT is nearly impossible with the comparator strategy, but EOT is cost-effective at a relatively low 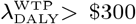, therefore yielding 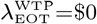 (Fig 1 and Table S2). The other regions present more complicated scenarios: EOT is likely in Region 2 (79%) and moderately likely in Region 3 (42%) even with the comparator strategy (Mean AS), but the value-for-money in terms of EOT objectives vary by location. In Region 2 when 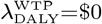, the Premium_EOT_=$229,000 and on a per-percentage-point basis 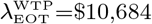 (Table S3). Region 3 has a moderate Premium_EOT_ of $651,000 for switching to a strategy sure to deliver EOT, but on a per-percentage-point basis, the cost 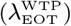 is $11,210 (Table S4). By contrast, although the Premium_EOT_ is highest in Region 1 ($709,800), it is the place where investments justified on the ground of elimination alone are the most efficient, 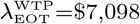. In short, while one could justify strategies that maximize the probability of elimination with a sufficiently high 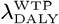 in Regions 1 and 2, that would not be possible in Region 3. In this way, if the current framework is employed simultaneously for various settings, as we have done here, or for multiple diseases marked for elimination, the results could aid in allocating resources to those locations or diseases where efforts for EOT are most efficient, starting where 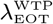 is lowest.

Existing literature on the economic evaluation of potential elimination strategies has been limited to diseases of person-to-person transmission, often considering one modality of prevention so that control vs elimination is a matter of the degree of coverage of vaccination (as with smallpox) or of treatment (as with HIV). This has made the analyses parsimonious, but it is unclear how such approaches generalize to real-world scenarios, where there are multiple modalities of control (3, 9, 10). In the case of gHAT as well as multiple other diseases, it is not clear that one activity alone is the key to elimination. Our framework therefore expands the categories of diseases that could be analyzed via a common set of metrics amenable to simulation analyses: 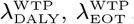, and Premium_EOT_. Alternatively, one could employ the same framework for another goal (i.e. reduction in inequality) to which there are benefits not captured by DALYs averted.

Our contention is that EOT is often not something that can be purchased outright, but rather something that can be invested on, and therefore the willingness-to-pay must be expressed in terms of the increased probability of reaching the goal. However, ascertaining the value of 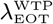 would probably be easier by asking stakeholders how much they would pay for elimination in a context where the probability of EOT is certain such that the probability is 0 or 1 with different strategies (like in region 1).

The EEOT of neglected tropical diseases is believed to drive towards achievement of several of the sustainable development goals (2, 33) as well as equity, as the elimination or eradication of a disease provides the same protection against that disease to individuals across the socioeconomic spectrum (34). Under the present framework, we contend that the investment on EEOT beyond its disease burden aspect (in terms of DALYs) are not only possible within campaigns that succeed, but that these benefits are achieved in relation to how close a strategy comes to elimination in terms of its probability of success. However, one does not necessarily need to enumerate each benefit of elimination and its relation to other social values, one only needs to have reasonable evidence of the presence of stakeholders that value EEOT to justify employing the approach we have presented here. Because the data to enumerate additional non-health benefits of elimination could be missing or weak, we propose an approach that circumvents such challenges by presenting the necessary lower bound for the willingness-to-pay for elimination.

### Limitations

The total costs of EEOT will inevitably be affected by the size of the population that must be treated, which involves the concepts of critical community size, the degree of connectivity of meta-populations, and importation probability–all factors that we do not examine here. Some diseases are worth eliminating in small patches because even one imported case will not re-establish transmission, while other diseases are only worth eliminating if elimination can be achieved throughout large interconnected networks of settlements (35–37).

Unlike many neglected tropical diseases, gHAT interventions have been very heterogeneous, even across the same administrative district, and so two regions with the same transmission in 2017 may have quite different underlying epidemiology. We captured this uncertainty by utilising posterior parameters from various regions, however the present results are not designed to be representative of a single area. Tailored models, fitted to longitudinal case and intervention data will yield more reliable location-specific recommendations for gHAT strategies.

It is worth noting that the premium of elimination is not a subsidy, although this number could be used to inform a subsidy. Incurring outlays in the short-term may require financing products even in the context of a strategy that could be cost-saving in the long run.

While we do not address issues surrounding the elimination of diseases and their inequitable distribution, there is a budding literature regarding such concerns termed ‘equity-enhanced CEA’, ‘extended CEA’, and ‘distributional CEA’ which look at how elimination and control can improve not only health but correct past inequities (32, 38–40).

## Conclusion

In conclusion, with this method, one can attest whether health-resource efficiency, in terms of DALYs, is enough to justify efforts that bring about elimination. The difference between the preferred strategy to reach each objective and the joint preferred strategy may set the basis for discussions on joint actions between stakeholders. In presenting such a flexible framework, it is tractable to perform analyses that could inform elimination priorities both within and across disease portfolios even when the benefits of elimination cannot be enumerated exactly.

## Materials and Methods

### An application: human African trypanosomiasis

*Gambiense* human African trypanosomiasis (gHAT) is a parasitic infection caused by *Trypanosoma brucei gambiense* and transmitted by tsetse (biting flies). gHAT infections are almost always fatal if untreated, and at the peak of the epidemic in the late 1990’s, it is suspected that up to tens of thousands of cases went undetected and untreated (41, 42). In 2012, the World Health Organization marked gHAT for elimination of transmission by 2030 (43). While gHAT has historically burdened 14 countries, the Democratic Republic of Congo (DRC) remains the most affected, accounting for over 74% of the worldwide caseload (44).

Here we employed a previously published model of gHAT transmission fitted to historic data from three health zones in DRC: Kwamouth, in Mai Ndombe province; Mosango, in Kwilu province; Sia, in Kwilu province. Details about these health zones are in SI section 1 and in a previous publication (45). Previously published models are based on epidemiological data provided by the WHO HAT Atlas (46). We selected these locations as they provide interesting illustrative examples of the NMEB framework.

### Health effects, costs, and cost-effectiveness

All modelling choices are described in previous publications (45, 47, 48) and summarised in the supplement (SI section 1). The model provided projections under alternative strategies for 2020–2040 of future case-reporting as well as unobservable features such as transmission events, disease burden, and unreported deaths (47). Four strategies made up of combinations of interventions are shown in Table 2 and illustrated in SI Fig S1.

We then applied a model of the resource use for these strategies (48) to estimate the costs and health burden accrued and averted in terms of cases, deaths, and DALYs. Costs were denominated in 2018 US$. Both costs and health effects are discounted at a rate of 3% in accordance with standard practice (20) and we performed our main analysis from the perspective of the healthcare providers collectively over a 20-year time-horizon (2020–2040).

Uncertainty was accounted for in two ways: 1) uncertainty in all model parameters was propagated via Monte Carlo simulation, drawing 10,000 random samples from probability distributions chosen to characterise the extant uncertainty in each parameter in accordance with established practice (23), and 2) the model-simulated stochasticity in case detections.

Because we are concerned with cost-effectiveness and uncertainty, we construct cost-effectiveness acceptability frontiers (CEAFs), which denote the optimal strategy (in terms of cost-effectiveness) at a range of willingness to pay values (14). Lastly, we develop the cost-effectiveness acceptability heatmaps (CEAHs), a form of two-way CEAF with both *λ* (WTP) values as x- and y-axis and the preferred intervention indicated by the color of the area of the heatmap. We use no pre-defined thresholds for WTP values, as we aim to provide guidance rather than prescription.

## Data Availability

All data and code to reproduce the analysis contained in this paper are available through OpenScienceFramework (OSF).

https://osf.io/fh6ca/

## Acknowledgements

We wish to thank Dr. Ron Crump for original epidemiological model fit to data (45), and Dr. Erick Miaka of the Programme National de Lutte contre la Trypanosomiase Humaine Africaine (PNLTHA) in DRC and Dr. Jose Ramón Franco at the World Health Organization (WHO) for providing the data (46) which was used for original model fitting (45) and ongoing discussions on gHAT with our group. This work was funded by the Bill and Melinda Gates Foundation (BMGF): Human African Trypanosomiasis Modelling and Economic Predictions for Policy (HAT MEPP) project (OPP1177824) and the NTD Modelling Consortium (OPP1184344, OPP1156227, and OPP1186851).

