## Supplemental Text 1 for "Economic evaluation of disease elimination: an extension to the net-benefit framework and application to human African trypanosomiasis"

PNAS
www.pnas.org

### Supplementary Information for

#### Economic evaluation of disease elimination: an extension to the net benefits framework and application to human African trypanosomiasis

**Marina Antillón, Ching-I Huang, Kat S Rock, Fabrizio Tediosi**

**Corresponding author: Marina Antillón.**
****

##### This PDF file includes:

Supplementary text
Figs. S1 to S5
Tables S1 to S4
SI References

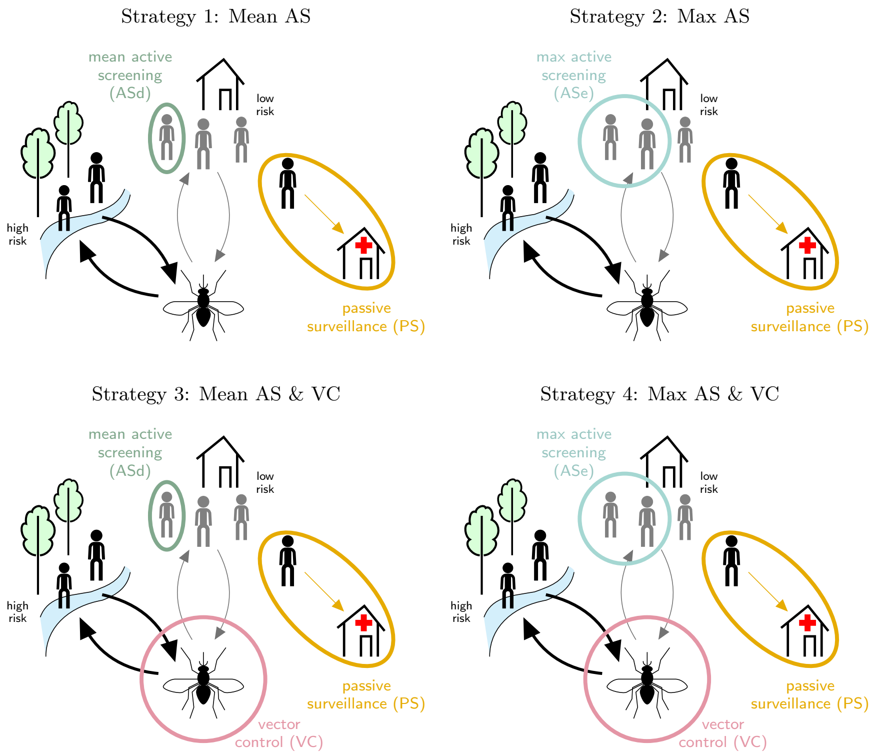


**Fig. S1.** Model of strategies against gHAT in DRC including active screening (AS) by mobile teams, passive surveillance (PS) in fixed health facilities. Cases detected by either mode are treated. In two strategies ('Mean AS' and 'Mean AS & VC') the active screening coverage is equal to the mean number screened during 2014–2018. In two other strategies ('Max AS' and 'Max AS & VC'), the coverage is the maximum number screened during 2000–2018. In strategies 3 and 4 vector control (VC) is simulated assuming a 80% tsetse density reduction after one year. PS is in place under all strategies. Figure reproduced under the creative commons licence from Antillon *et al.* (48).

### Supporting Information Text

#### 1. Supplementary methods

**A. Disease model.** We employed a previously published dynamic, deterministic transmission Susceptible-Exposed-Infected-Recovered-Suceptible (SEIRS) model. While the model is detailed elsewhere (45**?** ), briefly, we simulated gHAT illness and vector transmission in a compartmental model described by deterministic features, simulated using a set of ordinary differential equations. The stochastic nature of the observations was conferred by sampling from the infected prevalence to simulate those individuals who are diagnosed and report for treatment, either in a fixed health facility or to a mobile screening unit, and whether imperfect diagnostics correctly detect cases or identifies non-infected people as cases.

A Markov Chain Monte Carlo (MCMC) approach was previously used to generate posterior parameter sets for which the model outputs match the longitudinal data for different regions of the Democratic Republic of Congo (DRC) during 2000–2016. In the present study we selected three example regions – Kwamouth (Region 1), Mosango (Region 2), and Sia (Region 3) – of the 168 originally fitted in order to highlight diverse results of our new framework. The locations used in the current analysis are described in Table S1.

**Table S1. Descriptive summaries of three health zones.**

| Characteristic | Kwamouth | Mosango | Sia |
|---|---|---|---|
| Province | Mai-Ndombe | Kwilu | Kwilu |
| Population (2016 est.) | 131,022 | 125,076 | 114,041 |
| Area (km$^2$) | 14,589 | 2,606 | 2,990 |
| Active screening as a percent of 2016 population (mean; max) | 48; 69 | 34; 60 | 19; 30 |
| HAT testing centers (2014 est.) | 5 | 1 | 1 |
| Yearly incidence per 10,000 (2014–2018) | 8.70 | 0.99 | 1.02 |
| WHO Incidence category (2014–2018) | Moderate | Low | Moderate |
| Vector control extent (linear km) | 432 | 210 | 210 |
| Vector control density (targets per linear km) | 40 | 40 | 40 |

N.B.: For Kwamouth, the extent of riverbank where vector control must be performed is informed by planned activities. For Mosango and Sia, assumptions regarding vector control are based on the experience in places of similar size.

#### B. Strategies and intervention model.

***B.1. Medical Interventions.*** While the projections of strategies have been detailed in previously published manuscripts (47, 48), we provide a brief sketch of the features of the simulation within the context of the current analysis. To determine the number of cases detected by screening as well as the time lived with disease for cases that were never detected, we simulated a diagnostic algorithm combined with the prevalence determined by the transmission model. Although diagnostic algorithms are elaborate in practice, we simulated a simple algorithm that would capture the major features of the real process (**? ?** ). Suspects in traditional active or passive surveillance activities are first screened by the Card Agglutination tests for Trypanosomiasis (CATT) or rapid diagnostic tests (RDTs). Serologically-positive suspects then have blood drawn for microscopy, and if trypanosoma are found, a patient undergoes a lumbar puncture to stage their disease (early infection is "stage 1" and late is "stage 2") and determine care. For those ineligible for oral fexinidazole treatment, staging of the disease is done via lumbar puncture and followed by either treatment with pentamidine (stage 1) or nifurtimox-eflornithine combination treatment (NECT, stage 2) (26, 48).

***B.2. Vector control.*** To control the population of tsetse, special "Tiny Targets" have been developed that stand on riversides – typical tsetse habitat – and deliver deadly insecticide upon contact (27**? ? ?** ). The advantage of this method of control of disease is that it breaks the chain of transmission, even when some cases cannot be reached for treatment. Activities entail placing Tiny Targets alongside the riverbanks twice per year for as many years as it takes to see a decline in the transmission of cases. The impact of these activities on tsetse density has been documented elsewhere (27**?** ) and its impact on disease transmission has been evaluated in one modeling study for Chad (29) and through analysis of case reporting in Guinea (**?** ).

In our study, we calculate that vector control will have to be deployed along 437 km of riverbank in Kwamouth but only 210 km of riverbank in Mosango and Sia. The reason is that we assumed that smaller health zones like Mosango and Sia would need vector control activities closer to those in Yasa Bonga, a health zone of 2,606 km$^2$ (**?** ). Kwamouth, by contrast, spans 14,589 km$^2$ and contains two hot spots of transmission and therefore requires a broader treatment of riverbanks (48).

***B.3. Interventions in the endgame.*** In simulating the end-game, we also assumed that additional confirmatory procedures such as video microscopy (or lab-based tests) are being used to elevate the previous high specificity of screening algorithms ($\approx$ 99.9%) to 100% in this context of diminishing prevalence (45). We further simulate the impact of stopping active screening and vector control interventions (where applicable) based on observing three consecutive years of zero cases reporting (in either active screening or passive surveillance). Our algorithm would allow restarting active screening should further cases later arise through continuing passive detection. This cessation criterion is not only plausible in practice (it is unlikely that interventions would continue indefinitely) but also is important in capturing the impact of stopping transmission and therefore saving future intervention costs.

**C. Key outputs.** The key outputs of the dynamic and diagnostic models include mortality in undetected cases, detected cases in stages 1 and 2, and DALYs before and after presenting to care for all interventions. The number of people actively screened, and number (if any) of vector deployments performed each year is recorded.

**D. Elimination of Transmission.** Elimination of transmission (EOT) is assumed when the underlying transmission (not detected cases) falls below 1 new infection per year (this proxy threshold is necessary when using a deterministic model to approximate peri-elimination dynamics and has been used elsewhere (47**?** ). The metric of interest in this paper is regional EOT (where baseline activities must remain to prevent re-establishment) rather than eradication because we are not treating the issue of importation of cases (**?** ). This is also related to the World Health Organization's gHAT goal for 2030 which is global EOT to humans (43).

**E. Health impact.** We measured health impact by combining the epidemiological outputs from the dynamic/screening model with a probability tree that simulates the branching process of treatment progression and disease outcomes.

In accordance with the WHO interim guidelines on gHAT treatment (26), we assumed that treatment would consist of 1800 mg of fexinidazole for four days and 1200 mg of fexinidazole for six days for most patients in stage 1 and 2 disease. We assumed that patients would be treated in equal parts on an inpatient and outpatient basis (as directly-observed therapy). Patients who were either under 6 years old or under 20 kg in weight, undergo a lumbar puncture to determine disease stage and are sorted accordingly into pentamidine or NECT inpatient treatment.

We simulated the disease process separately for stage 1 and stage 2 disease, and a small proportion of stage 1 cases are assumed not cooperate with care or to undergo treatment failure, and are thus added to the number of cases that undergo stage 2 care.

The disease and treatment probability tree model is deterministic, formalized mathematically through the product of conditional probabilities of the outcome at each stage of disease and treatment progression (Fig S2). The disease tree model for stage 1 includes: follow-up (for patients lost-to-follow-up), the presence of side effects, treatment success or failure, diagnosis (in the case of treatment failure), and progression to stage 2 treatment (if applicable). For stage 2, additional steps include death due to treatment and the process of rescue treatment (for patients who fail first-choice treatment for stage 2).

Health burden is denominated in DALYs, but we report cases and deaths for the benefit of the reader (Table 3). The probability of EOT is denominated as a probability, and we treat it separately to DALYs averted.

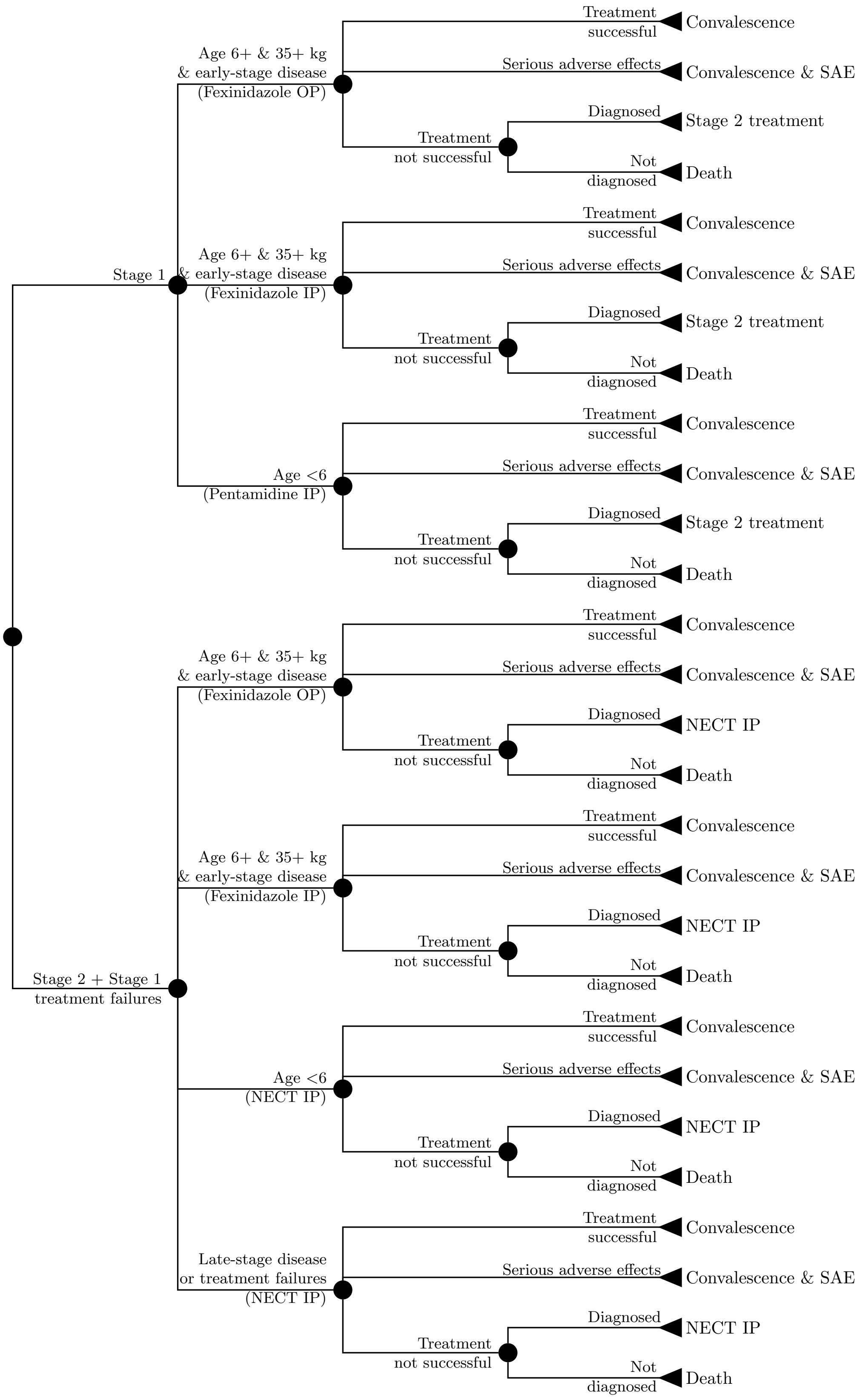


**Fig. S2.** Model of treatment for gHAT stages 1 and 2. Figure reproduced under the creative commons licence from Antillon $et\ al$ (48).

**F. Costs.** We performed our analysis using the health payer perspective, and therefore, only direct medical costs are included in our analysis. The cost function structure is identical to that described in (48), and therefore we only briefly describe the main features here, but assumptions behind consumption vary slightly.

Intervention costs are estimated as the product of unit costs and appropriate units as informed by the extant literature on interventions and our collaborators in DRC. Accelerating costs per case found towards the end-game is the results from a decreasing number of cases.

Disease costs are linked to the probability tree model of treatment progression, and include diagnosis, confirmation, and staging, as well as the cost of the drug itself and the administration. Costs values are drawn from the literature. Where no cost data existed, WHO CHOICE costs were used. All costs are inflated to 2018 US$ values.

As with the health impact, we compute costs using a 20-year time-horizon to allow time to consider the economic rewards of elimination by 2030. However, any other choice of time horizon would be equally amenable to use in this new framework.

#### G. Cost-effectiveness.

***G.1. Risk-averse vs risk-neutral decision-makers.*** The normative interpretation can be made in two ways: a risk-averse decision-maker will select the strategy that has the highest chance of being cost-effective; however, the risk-neutral decision-maker might not, due to an interesting feature in probabilistic simulations. One might note that in situations in which the probability distributions of the parameters in the simulation are asymmetric, some interventions might have a lower median than a mean, due to long tails in their distributions, thus sometimes yielding low probabilities of cost-effectiveness among all simulations but high expectations in the NMB. A risk-neutral decision-maker who looks only to maximize the health returns on a given WTP would choose the strategy with the highest expected NMB:

$$\underset{k \in 1:J}{\operatorname{argmax}} \, \mathbb{E}(\mathrm{NMB}(k, \theta_i | \lambda^{\mathrm{WTP}}))$$

That debate is beyond the scope of the paper, and we will proceed with an assumption of a risk-neutral decision-maker.

***G.2. Time horizons.*** We chose a 20-year time-horizon starting in 2020 to allow the economic rewards of elimination by 2030 to be reaped.

#### 2. Supplemental results

**Table S2. Intermediate outcomes, cost-effectiveness, and efficiency of elimination in Region 1.**

| | Mean AS | Max AS | Mean AS & VC | Max AS & VC |
|---|---|---|---|---|
| **Basic outputs** | | | | |
| Cases | 477 (144, 1,081) | 463 (136, 1,047) | 116 (41, 235) | 120 (38, 270) |
| Deaths | 207 (41, 614) | 174 (36, 499) | 54 (18, 115) | 49 (16, 105) |
| DALYs | 3,939 (886, 11,007) | 3,336 (779, 9,161) | 1,185 (405, 2,494) | 1,077 (362, 2,280) |
| $\Delta$DALYs | Comparator | 602 (-191, 2,221) | 2,754 (339, 8,765) | 2,862 (382, 8,956) |
| Costs (USD, $\times$ 1000) | 3,101 (2,153, 4,736) | 4,023 (2,734, 6,308) | 3,811 (2,464, 6,007) | 4,284 (2,732, 6,731) |
| $\Delta$Costs (USD, $\times$ 1000) | Comparator | 921 (451, 1,619) | 709.8 (-763.9, 2,765) | 1,182 (-291.7, 3,415) |
| ICER | Minimum cost | Dominated | 258 | 4,373 |
| Pr. EOT | 0 | 0 | 100 | 100 |
| $\Delta$Pr. EOT | Comparator | 0 | 100 | 100 |
| **$\lambda^{WTP}_{DALY}$ = 0 USD** | | | | |
| NMB (USD, $\times$ 1000) | 0 (0, 0) | -921.2 (-1,619, -450.7) | -709.8 (-2,765, 763.9) | -1,182 (-3,415, 291.7) |
| Justifiable costs (USD, $\times$ 1000) | Preferred | 0 (0, 0) | 0 (0, 0) | 0 (0, 0) |
| $Premium_{EOT}$ (USD, $\times$ 1000) | Preferred | 921 (451, 1,619) | 710 (0, 2,765) | 1,182 (0, 3,415) |
| $\Delta Premium_{EOT}/\Delta$Pr. EOT (vs Preferred, USD) | Preferred | No advantage | 7,098 | 11,821 |
| $\Delta Premium_{EOT}/\Delta$Pr. EOT (incremental, USD) | Preferred | Dominated | 7,098 | Dominated |
| **$\lambda^{WTP}_{DALY}$ = 250 USD** | | | | |
| NMB (USD, $\times$ 1000) | 0 (0, 0) | -770.6 (-1,501, -211.2) | -21.37 (-2,486, 2,287) | -466.7 (-3,096, 1,838) |
| Justifiable costs (USD, $\times$ 1000) | Preferred | 150.6 (-47.86, 555.2) | 688 (85, 2,191) | 715 (95, 2,239) |
| $Premium_{EOT}$ (USD, $\times$ 1000) | Preferred | 771 (211, 1,501) | 21 (0, 2,486) | 467 (0, 3,096) |
| $\Delta Premium_{EOT}/\Delta$Pr. EOT (vs Preferred, USD) | Preferred | No advantage | 214 | 4,667 |
| $\Delta Premium_{EOT}/\Delta$Pr. EOT (incremental, USD) | Preferred | Dominated | 214 | Dominated |
| **$\lambda^{WTP}_{DALY}$ = 500 USD** | | | | |
| NMB (USD, $\times$ 1000) | 0 (0, 0) | -620 (-1,430, 243.9) | 667 (-2,248, 4,273) | 248.7 (-2,845, 3,910) |
| Justifiable costs (USD, $\times$ 1000) | Suboptimal | Suboptimal | Preferred | 54.01 (-146.8, 280) |
| $Premium_{EOT}$ (USD, $\times$ 1000) | Suboptimal | Suboptimal | Preferred | 418 (0, 1,839) |
| $\Delta Premium_{EOT}/\Delta$Pr. EOT (vs Preferred, USD) | Suboptimal | Suboptimal | Preferred | No advantage |
| $\Delta Premium_{EOT}/\Delta$Pr. EOT (incremental, USD) | Suboptimal | Suboptimal | Preferred | Dominated |
| **$\lambda^{WTP}_{DALY}$ = 1000 USD** | | | | |
| NMB (USD, $\times$ 1000) | 0 (0, 0) | -318.9 (-1,379, 1,296) | 2,044 (-1,813, 8,517) | 1,680 (-2,418, 8,327) |
| Justifiable costs (USD, $\times$ 1000) | Suboptimal | Suboptimal | Preferred | 108 (-293.6, 560.1) |
| $Premium_{EOT}$ (USD, $\times$ 1000) | Suboptimal | Suboptimal | Preferred | 364 (0, 1,825) |
| $\Delta Premium_{EOT}/\Delta$Pr. EOT (vs Preferred, USD) | Suboptimal | Suboptimal | Preferred | No advantage |
| $\Delta Premium_{EOT}/\Delta$Pr. EOT (incremental, USD) | Suboptimal | Suboptimal | Preferred | Dominated |
| **$\lambda^{WTP}_{DALY}$ = 1500 USD** | | | | |
| NMB (USD, $\times$ 1000) | 0 (0, 0) | -17.69 (-1,404, 2,406) | 3,421 (-1,469, 12,859) | 3,110 (-2,036, 12,765) |
| Justifiable costs (USD, $\times$ 1000) | Suboptimal | Suboptimal | Preferred | 162 (-440.4, 840.1) |
| $Premium_{EOT}$ (USD, $\times$ 1000) | Suboptimal | Suboptimal | Preferred | 310 (0, 1,846) |
| $\Delta Premium_{EOT}/\Delta$Pr. EOT (vs Preferred, USD) | Suboptimal | Suboptimal | Preferred | No advantage |
| $\Delta Premium_{EOT}/\Delta$Pr. EOT (incremental, USD) | Suboptimal | Suboptimal | Preferred | Dominated |

[1] A dominated strategy is one that has a higher cost but averts fewer DALYs or has the same or lower probability of EOT than a less expensive strategy.
[2] We do not show prediction intervals for ICERs as there are a variety of issues with the mathematical properties of such constructions (21).
[3] For context on the values of $\lambda^{WTP}_{DALY}$ = that we have chosen to display, see Table 2.

**Table S3. Intermediate outcomes, cost-effectiveness, and efficiency of elimination in Region 2.**

| | Mean AS | Max AS | Mean AS & VC | Max AS & VC |
|---|---|---|---|---|
| **Basic outputs** | | | | |
| Cases | 23 (1, 79) | 22 (0, 92) | 9 (0, 41) | 10 (0, 54) |
| Deaths | 12 (1, 42) | 8 (0, 28) | 5 (0, 15) | 4 (0, 12) |
| DALYs | 247 (20, 803) | 167 (2, 564) | 106 (1, 318) | 82 (1, 262) |
| $\Delta$DALYs | Comparator | 80 (-87, 366) | 142 (-41, 551) | 165 (-21, 597) |
| Costs (USD, $\times$ 1000) | 1,029 (508, 1,841) | 1,407 (637, 2,652) | 1,258 (636, 2,068) | 1,529 (743, 2,544) |
| $\Delta$Costs (USD, $\times$ 1000) | Comparator | 377.5 (-164.3, 1,105) | 229 (-451.9, 933.8) | 499.7 (-209.8, 1,335) |
| ICER | Minimum cost | Dominated | 1,615 | 11,578 |
| Pr. EOT | 79 | 92 | 100 | 100 |
| $\Delta$Pr. EOT | Comparator | 13 | 21 | 21 |
| $\lambda^{\mathrm{WTP}}_{\mathrm{DALY}}$ **= 0 USD** | | | | |
| NMB (USD, $\times$ 1000) | 0 (0, 0) | -377.5 (-1,105, 164.3) | -229 (-933.8, 451.9) | -499.7 (-1,335, 209.8) |
| Justifiable costs (USD, $\times$ 1000) | Preferred | 0 (0, 0) | 0 (0, 0) | 0 (0, 0) |
| $\text{Premium}_{\text{EOT}}$ (USD, $\times$ 1000) | Preferred | 377 (0, 1,105) | 229 (0, 934) | 500 (0, 1,335) |
| $\Delta\text{Premium}_{\text{EOT}}/\Delta$Pr. EOT (vs Preferred, USD) | Preferred | 29,126 | 10,684 | 23,318 |
| $\Delta\text{Premium}_{\text{EOT}}/\Delta$Pr. EOT (incremental, USD) | Preferred | Dominated | 10,684 | Dominated |
| $\lambda^{\mathrm{WTP}}_{\mathrm{DALY}}$ **= 250 USD** | | | | |
| NMB (USD, $\times$ 1000) | 0 (0, 0) | -357.5 (-1,073, 177) | -193.5 (-912.2, 515.7) | -458.4 (-1,300, 271.7) |
| Justifiable costs (USD, $\times$ 1000) | Preferred | 20 (-21.73, 91.53) | 35.43 (-10.22, 137.7) | 41.28 (-5.223, 149.3) |
| $\text{Premium}_{\text{EOT}}$ (USD, $\times$ 1000) | Preferred | 357 (0, 1,073) | 194 (0, 912) | 458 (0, 1,300) |
| $\Delta\text{Premium}_{\text{EOT}}/\Delta$Pr. EOT (vs Preferred, USD) | Preferred | 27,582 | 9,031 | 21,391 |
| $\Delta\text{Premium}_{\text{EOT}}/\Delta$Pr. EOT (incremental, USD) | Preferred | Dominated | 9,031 | Dominated |
| $\lambda^{\mathrm{WTP}}_{\mathrm{DALY}}$ **= 500 USD** | | | | |
| NMB (USD, $\times$ 1000) | 0 (0, 0) | -337.5 (-1,048, 189.1) | -158.1 (-889.4, 593.1) | -417.1 (-1,269, 339.8) |
| Justifiable costs (USD, $\times$ 1000) | Preferred | 40.01 (-43.47, 183.1) | 70.87 (-20.45, 275.4) | 82.56 (-10.45, 298.7) |
| $\text{Premium}_{\text{EOT}}$ (USD, $\times$ 1000) | Preferred | 337 (0, 1,048) | 158 (0, 889) | 417 (0, 1,269) |
| $\Delta\text{Premium}_{\text{EOT}}/\Delta$Pr. EOT (vs Preferred, USD) | Preferred | 26,039 | 7,377 | 19,465 |
| $\Delta\text{Premium}_{\text{EOT}}/\Delta$Pr. EOT (incremental, USD) | Preferred | Dominated | 7,377 | Dominated |
| $\lambda^{\mathrm{WTP}}_{\mathrm{DALY}}$ **= 1000 USD** | | | | |
| NMB (USD, $\times$ 1000) | 0 (0, 0) | -297.4 (-1,013, 235.9) | -87.23 (-854.9, 779.6) | -334.6 (-1,225, 533.6) |
| Justifiable costs (USD, $\times$ 1000) | Preferred | 80.02 (-86.94, 366.1) | 141.7 (-40.89, 550.7) | 165.1 (-20.89, 597.3) |
| $\text{Premium}_{\text{EOT}}$ (USD, $\times$ 1000) | Preferred | 297 (0, 1,013) | 87 (0, 855) | 335 (0, 1,225) |
| $\Delta\text{Premium}_{\text{EOT}}/\Delta$Pr. EOT (vs Preferred, USD) | Preferred | 22,951 | 4,070 | 15,613 |
| $\Delta\text{Premium}_{\text{EOT}}/\Delta$Pr. EOT (incremental, USD) | Preferred | Dominated | 4,070 | Dominated |
| $\lambda^{\mathrm{WTP}}_{\mathrm{DALY}}$ **= 1500 USD** | | | | |
| NMB (USD, $\times$ 1000) | 0 (0, 0) | -257.4 (-977.1, 312) | -16.36 (-817.6, 1,003) | -252 (-1,186, 766.8) |
| Justifiable costs (USD, $\times$ 1000) | Preferred | 120 (-130.4, 549.2) | 212.6 (-61.34, 826) | 247.7 (-31.34, 896) |
| $\text{Premium}_{\text{EOT}}$ (USD, $\times$ 1000) | Preferred | 257 (0, 977) | 16 (0, 818) | 252 (0, 1,186) |
| $\Delta\text{Premium}_{\text{EOT}}/\Delta$Pr. EOT (vs Preferred, USD) | Preferred | 19,864 | 764 | 11,760 |
| $\Delta\text{Premium}_{\text{EOT}}/\Delta$Pr. EOT (incremental, USD) | Preferred | Dominated | 764 | Dominated |

[1] A dominated strategy is one that has a higher cost but averts fewer DALYs or has the same or lower probability of EOT than a less expensive strategy.
[2] We do not show prediction intervals for ICERs as there are a variety of issues with the mathematical properties of such constructions (21).
[3] For context on the values of $\lambda^{\mathrm{WTP}}_{\mathrm{DALY}}$ = that we have chosen to display, see Table 2.

**Table S4. Intermediate outcomes, cost-effectiveness, and efficiency of elimination in Region 3.**

| | Mean AS | Max AS | Mean AS & VC | Max AS & VC |
|---|---|---|---|---|
| **Basic outputs** | | | | |
| Cases | 65 (2, 224) | 64 (1, 264) | 27 (1, 84) | 31 (0, 122) |
| Deaths | 32 (1, 137) | 19 (0, 89) | 14 (0, 54) | 10 (0, 44) |
| DALYs | 676 (23, 2,809) | 414 (4, 1,885) | 336 (10, 1,245) | 242 (3, 1,008) |
| $\Delta$DALYs | Comparator | 262 (-38, 1,133) | 340 (-50, 1,684) | 434 (-14, 1,926) |
| Costs (USD, $\times$ 1000) | 970 (524, 1,552) | 1,164 (573, 2,058) | 1,622 (869, 2,793) | 1,659 (882, 3,023) |
| $\Delta$Costs (USD, $\times$ 1000) | Comparator | 193.5 (-138.4, 599.4) | 651 (16, 1,613) | 689 (38, 1,763) |
| ICER | Minimum cost | 740 | Weakly dominated | 2,875 |
| Pr. EOT | 42 | 54 | 100 | 100 |
| $\Delta$Pr. EOT | Comparator | 12 | 58 | 58 |
| **$\lambda^{WTP}_{DALY}$ = 0 USD** | | | | |
| NMB (USD, $\times$ 1000) | 0 (0, 0) | -193.5 (-599.4, 138.4) | -651.4 (-1,613, -15.83) | -689 (-1,763, -37.58) |
| Justifiable costs (USD, $\times$ 1000) | Preferred | 0 (0, 0) | 0 (0, 0) | 0 (0, 0) |
| $\text{Premium}_{EOT}$ (USD, $\times$ 1000) | Preferred | 194 (0, 599) | 651 (16, 1,613) | 689 (38, 1,763) |
| $\Delta\text{Premium}_{EOT}/\Delta$Pr. EOT (vs Preferred, USD) | Preferred | 15,747 | 11,210 | 11,858 |
| $\Delta\text{Premium}_{EOT}/\Delta$Pr. EOT (incremental, USD) | Preferred | Weakly Dominated | 11,210 | Dominated |
| **$\lambda^{WTP}_{DALY}$ = 250 USD** | | | | |
| NMB (USD, $\times$ 1000) | 0 (0, 0) | -128.1 (-505.5, 180.6) | -566.4 (-1,507, 121.2) | -580.5 (-1,602, 141.1) |
| Justifiable costs (USD, $\times$ 1000) | Preferred | 65.4 (-9.513, 283.2) | 84.99 (-12.61, 421.1) | 108.5 (-3.581, 481.5) |
| $\text{Premium}_{EOT}$ (USD, $\times$ 1000) | Preferred | 128 (0, 505) | 566 (0, 1,507) | 581 (0, 1,602) |
| $\Delta\text{Premium}_{EOT}/\Delta$Pr. EOT (vs Preferred, USD) | Preferred | 10,425 | 9,747 | 9,991 |
| $\Delta\text{Premium}_{EOT}/\Delta$Pr. EOT (incremental, USD) | Preferred | Weakly Dominated | 9,747 | Dominated |
| **$\lambda^{WTP}_{DALY}$ = 500 USD** | | | | |
| NMB (USD, $\times$ 1000) | 0 (0, 0) | -62.72 (-449, 325.5) | -481.4 (-1,416, 383.2) | -472.1 (-1,476, 431.8) |
| Justifiable costs (USD, $\times$ 1000) | Preferred | 130.8 (-19.03, 566.4) | 170 (-25.21, 842.2) | 217 (-7.162, 963.1) |
| $\text{Premium}_{EOT}$ (USD, $\times$ 1000) | Preferred | 63 (0, 449) | 481 (0, 1,416) | 472 (0, 1,476) |
| $\Delta\text{Premium}_{EOT}/\Delta$Pr. EOT (vs Preferred, USD) | Preferred | 5,103 | 8,285 | 8,123 |
| $\Delta\text{Premium}_{EOT}/\Delta$Pr. EOT (incremental, USD) | Preferred | 5,103 | Dominated | 8,934 |
| **$\lambda^{WTP}_{DALY}$ = 1000 USD** | | | | |
| NMB (USD, $\times$ 1000) | 0 (0, 0) | 68.09 (-394.3, 789.6) | -311.4 (-1,304, 1,036) | -255.1 (-1,295, 1,237) |
| Justifiable costs (USD, $\times$ 1000) | Suboptimal | Preferred | 78.35 (-149.3, 689.6) | 172.4 (-62.55, 933.8) |
| $\text{Premium}_{EOT}$ (USD, $\times$ 1000) | Suboptimal | Preferred | 380 (0, 1,307) | 323 (0, 1,274) |
| $\Delta\text{Premium}_{EOT}/\Delta$Pr. EOT (vs Preferred, USD) | Suboptimal | Preferred | 8,283 | 7,053 |
| $\Delta\text{Premium}_{EOT}/\Delta$Pr. EOT (incremental, USD) | Suboptimal | Preferred | Dominated | 7,053 |
| **$\lambda^{WTP}_{DALY}$ = 1500 USD** | | | | |
| NMB (USD, $\times$ 1000) | 0 (0, 0) | 198.9 (-365.1, 1,352) | -141.5 (-1,234, 1,792) | -38.07 (-1,204, 2,134) |
| Justifiable costs (USD, $\times$ 1000) | Suboptimal | Preferred | 117.5 (-224, 1,034) | 258.5 (-93.82, 1,401) |
| $\text{Premium}_{EOT}$ (USD, $\times$ 1000) | Suboptimal | Preferred | 340 (0, 1,305) | 237 (0, 1,235) |
| $\Delta\text{Premium}_{EOT}/\Delta$Pr. EOT (vs Preferred, USD) | Suboptimal | Preferred | 7,428 | 5,172 |
| $\Delta\text{Premium}_{EOT}/\Delta$Pr. EOT (incremental, USD) | Suboptimal | Preferred | Dominated | 5,172 |

[1] A dominated strategy is one that has a higher cost but averts fewer DALYs or has the same or lower probability of EOT than a less expensive strategy.
[2] We do not show prediction intervals for ICERs as there are a variety of issues with the mathematical properties of such constructions (21).
[3] For context on the values of $\lambda^{WTP}_{DALY}$ = that we have chosen to display, see Table 2.

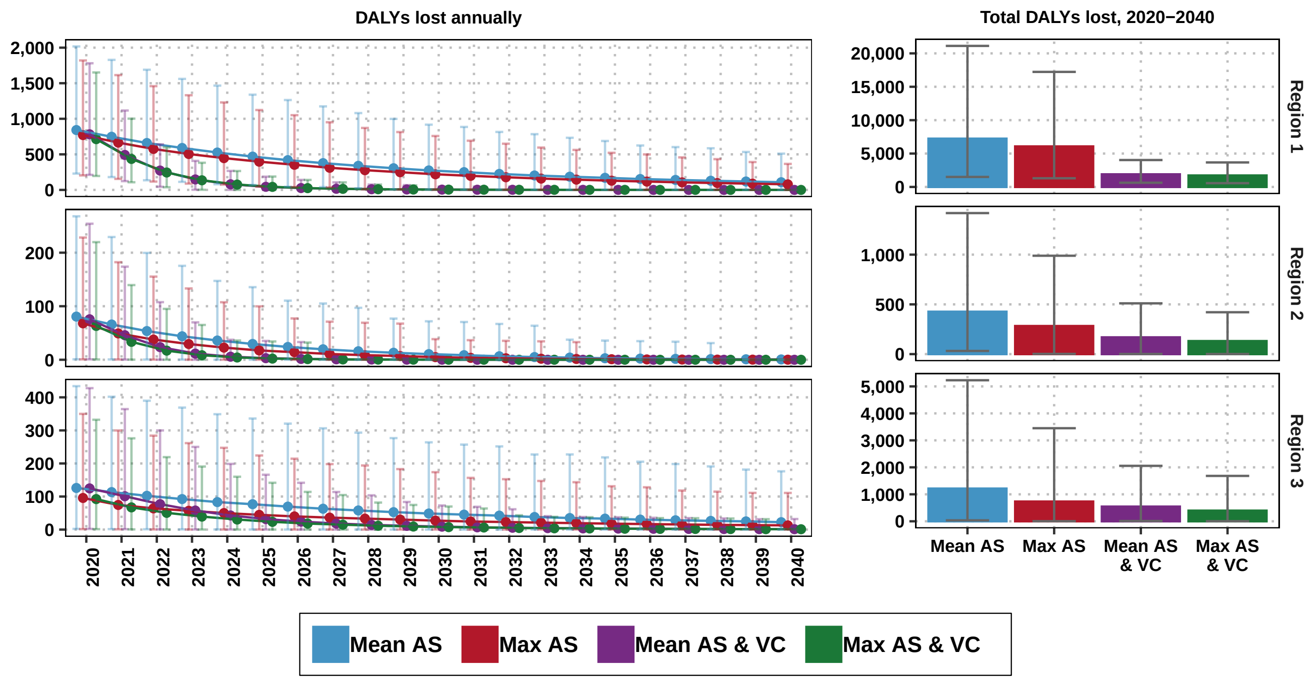


**Fig. S3.** Components of mean annual and cumulative costs, by strategy and location. Displayed costs are not discounted. Treatment costs, indicated in purple, are shown here although they are so small as to be hardly visible.

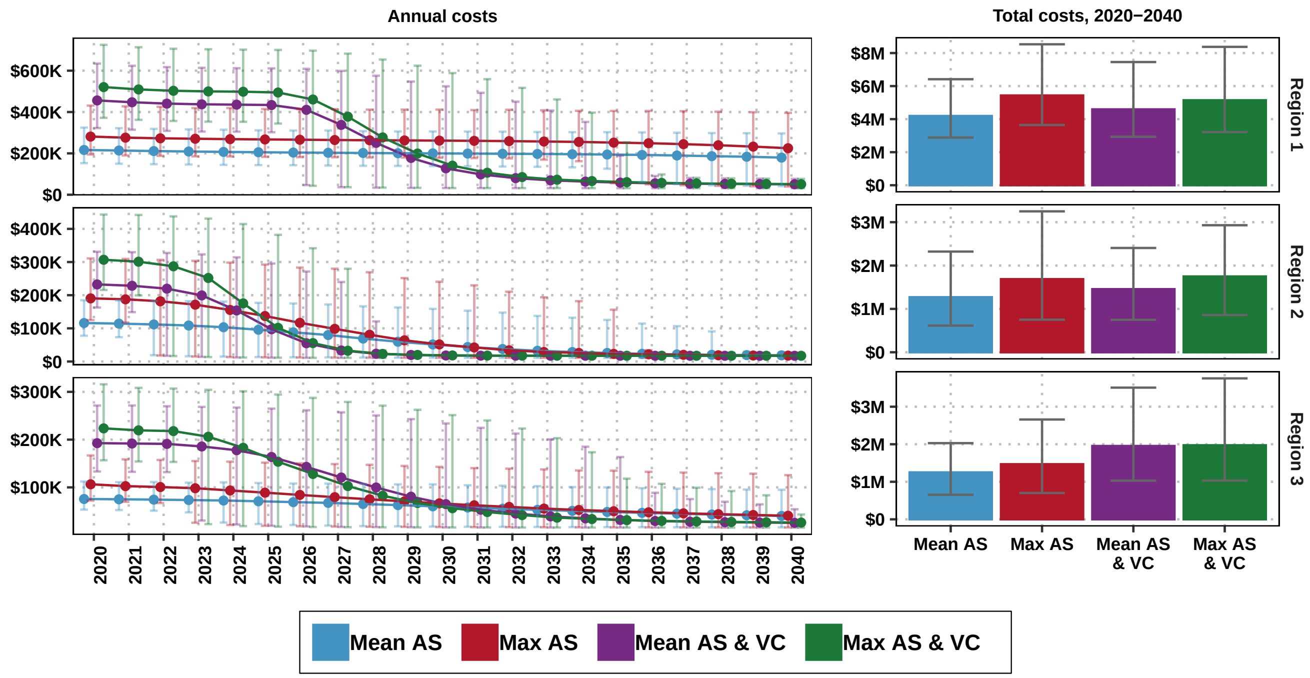


**Fig. S4.** Disability-adjusted life-years (DALYs) lost per strategy by location, 2020-2040. DALYs are not discounted. Estimates shown are means and their 95% predictive intervals (PI).

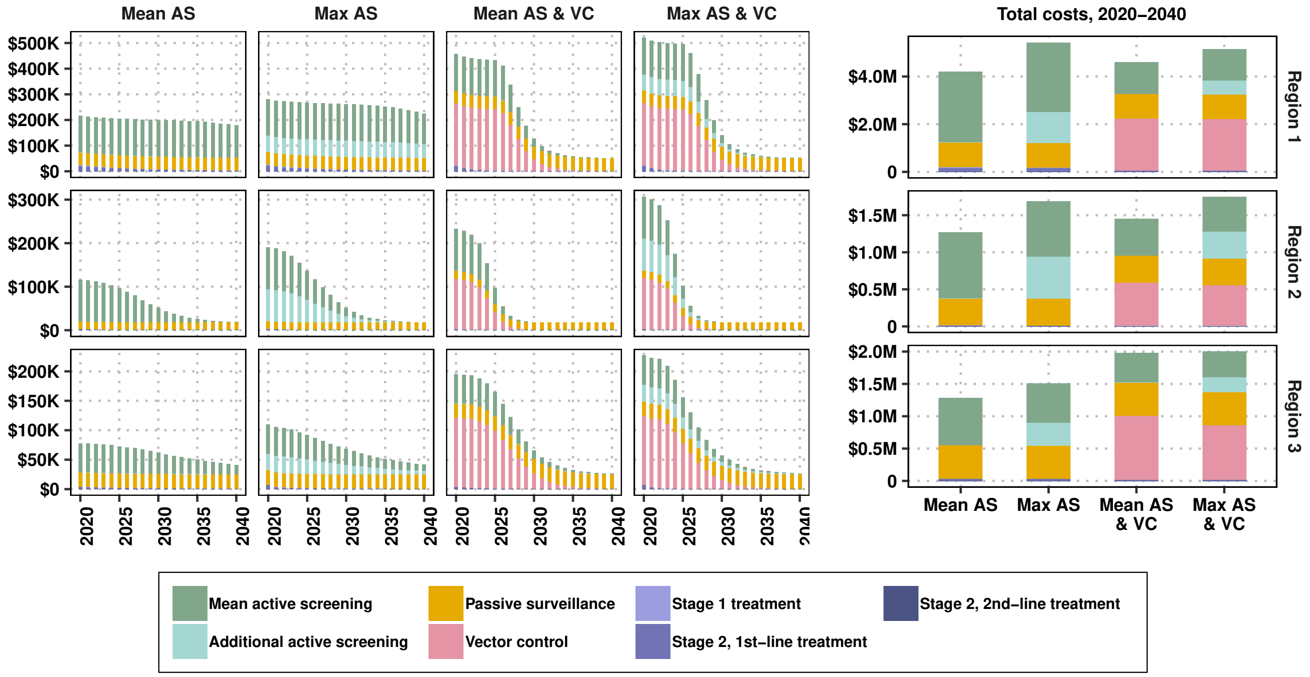


**Fig. S5.** Costs per strategy by location, 2020-2040. Costs are not discounted. Estimates shown are means and their 95% predictive intervals (PI).
